# Normative vs. patient-specific brain connectivity in Deep Brain Stimulation

**DOI:** 10.1101/2020.02.24.20027490

**Authors:** Qiang Wang, Harith Akram, Muthuraman Muthuraman, Gabriel Gonzalez-Escamilla, Sameer A. Sheth, Sergiu Groppa, Nora Vanegas-Arroyave, Ludvic Zrinzo, Ningfei Li, Andrea Kühn, Andreas Horn

**Affiliations:** Movement Disorders & Neuromodulation Unit, Department for Neurology, Charité - University Medicine Berlin, Germany; Unit of Functional Neurosurgery, UCL Queen Square Institute of Neurology, Queen Square, London WC1N 3BG, UK; Victor Horsley Department of Neurosurgery, National Hospital for Neurology and Neurosurgery, UCLH, Queen Square, London WC1N 3BG, UK; Movement Disorders and Neurostimulation, Biomedical Statistics and Mulitmodal Signal Processing Unit, Department of Neurology, University Medical Center of the Johannes Gutenberg University, Mainz, Germany; Department of Neurosurgery, Baylor College of Medicine, Houston, Texas, USA; Department of Neurology, Columbia University College of Physicians and Surgeons New York, NY, USA

**Keywords:** deep brain stimulation, subthalamic nucleus, Parkinson’s disease, human connectome, tractography

## Abstract

Brain connectivity profiles seeding from deep brain stimulation (DBS) electrodes have emerged as informative tools to estimate outcome variability across DBS patients. Given the limitations of acquiring and processing patient-specific diffusion-weighted imaging data, most studies have employed normative atlases of the human connectome. To date, it remains unclear whether patient-specific connectivity information would strengthen the accuracy of such analyses. Here, we compared similarities and differences between patient-specific, disease-matched and normative structural connectivity data and retrospective estimation of clinical improvement that they may generate.

Data from 33 patients suffering from Parkinson’s Disease who underwent surgery at three different centers were retrospectively collected. Stimulation-dependent connectivity profiles seeding from active contacts were estimated using three modalities, namely either patient-specific diffusion-MRI data, disease-matched or normative group connectome data (acquired in healthy young subjects). Based on these profiles, models of optimal connectivity were constructed and used to retrospectively estimate the clinical improvement in out of sample data.

All three modalities resulted in highly similar optimal connectivity profiles that could largely reproduce findings from prior research based on a novel multi-center cohort. Connectivity estimates seeding from electrodes when using either patient-specific or normative connectomes correlated significantly to primary motor cortex (R = 0.57, p = 0.001, R=0.73, p=0.001), supplementary motor area (R = 0.40, p = 0.005, R = 0.43, p = 0.003), pre-supplementary motor area (R = 0.33, p = 0.022, R = 0.33, p = 0.031), but not to more frontal regions such as the dorsomedial prefrontal cortex (R = 0.21, p = 0.17, R = 0.18, p = 0.17).

However, in a data-driven approach that estimated optimal whole-brain connectivity profiles, out-of-sample estimation of clinical improvements were made and ranged within a similar magnitude when applying either of the three modalities (R = 0.43 at p = 0.001 for patient-specific connectivity; R = 0.25, p = 0.048 for the age- and disease-matched group connectome; R = 0.31 at p = 0.028 for healthy-/young connectome).

**Conclusions:** The use of patient-specific connectivity and normative connectomes lead to identical main conclusions about which brain areas are associated with clinical improvement. Still, although results were not significantly different, they hint at the fact that patient-specific connectivity may bear the potential of estimating slightly more variance when compared to group connectomes. Our findings further support the role of DBS electrode connectivity profiles as a promising method to guide surgical targeting and DBS programming.

## Introduction

Deep brain stimulation (DBS) is a well-established treatment for Parkinson’s disease (PD), alleviating motor symptoms and improving quality of life (Deuschl et al., 2006; Schuepbach et al., 2013). DBS does not only exert focal effects (i.e. at the subthalamic nucleus; STN) but also affects distributed basal-ganglia cortical cerebellar networks (Accolla et al., 2016; Helmich et al., 2012; Horn et al., 2017b; Horn, 2019; Kahan et al., 2019; Lozano and Lipsman, 2013; Muthuraman et al., 2018). The impact of DBS on pathological networks has been highlighted in PD (Accolla et al., 2016; Horn, 2019; Kahan et al., 2019) as well as other diseases such as Essential Tremor (Al-Fatly et al., 2019), dystonia (Corp et al., 2019) and obsessive-compulsive disorder (J. C. Baldermann et al., 2019).

Using preoperative diffusion-weighted imaging (dMRI), Vanegas-Arroyave and colleagues assessed the connectivity patterns of clinically beneficial DBS electrodes in PD patients. Their results suggested that modulation of white matter tracts directed to the superior frontal gyrus and the thalamus were associated with effective DBS outcomes (Vanegas-Arroyave et al., 2016). Similarly, Akram and colleagues used preoperative diffusion-weighted imaging (dMRI) data to investigate the cortical connectivity patterns associated with treatment efficacy (Akram et al., 2017). Maximal improvement in cardinal motor symptoms was associated with connectivity of DBS electrodes to different cortical regions: tremor control with connectivity to primary motor cortex (M1), bradykinesia with the supplementary motor area (SMA) and rigidity to both prefrontal cortex (PFC) and SMA. These two studies acquired dMRI in each patient preoperatively. While this approach represents the gold-standard of practice, a practical limitation is that resulting cohort sizes will often be small, studies costly and pooling across centers non-straightforward due to inconsistencies in MR acquisition protocols. There is, however, a large fraction of DBS patients in whom no preoperative dMRI data were obtained and therefore the use of individual brain connectivity measures is not feasible. This is especially relevant in novel indications such as Alzheimer’s Disease (Baldermann et al., 2018; Ponce et al., 2016) or psychiatric indications (Hamani et al., 2011; Huys et al., 2019) where limited numbers of patients undergo surgery world-wide. The same applies to “classical diseases” (such as dystonia) that are treated with unconventional targets (such as the STN) (Ostrem et al., 2011; Yao et al., 2019). Other methods are required to explore connectivity in patient cohorts that lack individualized dMRI data since this is an opportunity to better understand the effects of DBS on the brain and to advance clinical care.

An alternative approach is the use of normative connectomes – i.e. atlases of average brain connectivity calculated from large cohorts of subjects (Ewert et al., 2018; Horn et al., 2014a, 2019; Horn and Blankenburg, 2016; Marek et al., 2011; Thomas Yeo et al., 2011; Yeh et al., 2018; Yeh and Tseng, 2011). A first study that explored this concept investigated functional and structural connectivity profiles of the ventral intermediate nucleus of the thalamus (Horn et al., 2017a). A second study then investigated optimal connectivity profiles for STN-DBS (Horn et al., 2017b). In this study, the optimal connectivity profiles were estimated on one cohort from a first DBS center and - by using this model - predicted the motor outcome in patients operated at a different DBS center (Horn et al., 2017b). Specifically, structural and functional connectivity between DBS electrodes and other brain regions were correlated with UPDRS-III changes across patients. This resulted in a connectivity ‘fingerprint’ of effective DBS electrodes. To validate these maps, similarity indices between each electrode’s connectivity profile from an independent cohort and the ‘optimal’ fingerprint were calculated. These were then used to estimate variability in clinical improvement.

This concept has since then been applied to explore connectivity associated with clinical or behavioral changes in multiple diseases (Al-Fatly et al., 2019; J. C. Baldermann et al., 2019; Johnson et al., 2019; J. Baldermann et al., 2019; de Almeida Marcelino et al., 2019; Irmen et al., 2019; Li et al., 2019; Neumann et al., 2018). With the increasing popularity of this approach, it is timely to compare the results achieved by examining patient-specific connectivity with those obtained when using normative connectivity data from other cohorts. One main limitation of the approach is that connectivity data taken from connectome atlases can never represent individual differences in connectivity profiles from the actual DBS patients of study. Thus, the use of connectome atlases has clear similarities to the use of other atlases. For instance, histological atlas information was applied to inform DBS for decades (Schaltenbrand G, 1977; Talairach and Tournoux, 1988). Similarly, subcortical atlases – for instance of the STN – have been widely applied to study DBS electrode placement (for an overview see (Ewert et al., 2018)). However, despite the conceptual similarities to other atlases, some aspects of connectome atlases are novel and require further study.

Here, we have retrospectively analyzed individual connectivity estimates that were based on 33 DBS patients’ diffusion imaging data (dMRI). We reproduced workflows that were previously published using normative datasets. Furthermore, we compared the extent of clinical improvement that could be estimated by using individualized dMRI data versus the use of two connectome atlases that were based on either healthy subjects or PD patients. By doing so, we explored the specific similarities and differences between these types of connectivity information when seeding from DBS electrodes to the rest of the brain.

## Materials and methods

### Patient cohorts and imaging

Thirty-three DBS patients from 3 different DBS centers (Center 1 (London): N = 17, Center 2 (Mainz): N = 12, Center 3 (New York): N = 4) were included in this retrospective study. Patient demographics are summarized in Table 1.

**Table 1.**
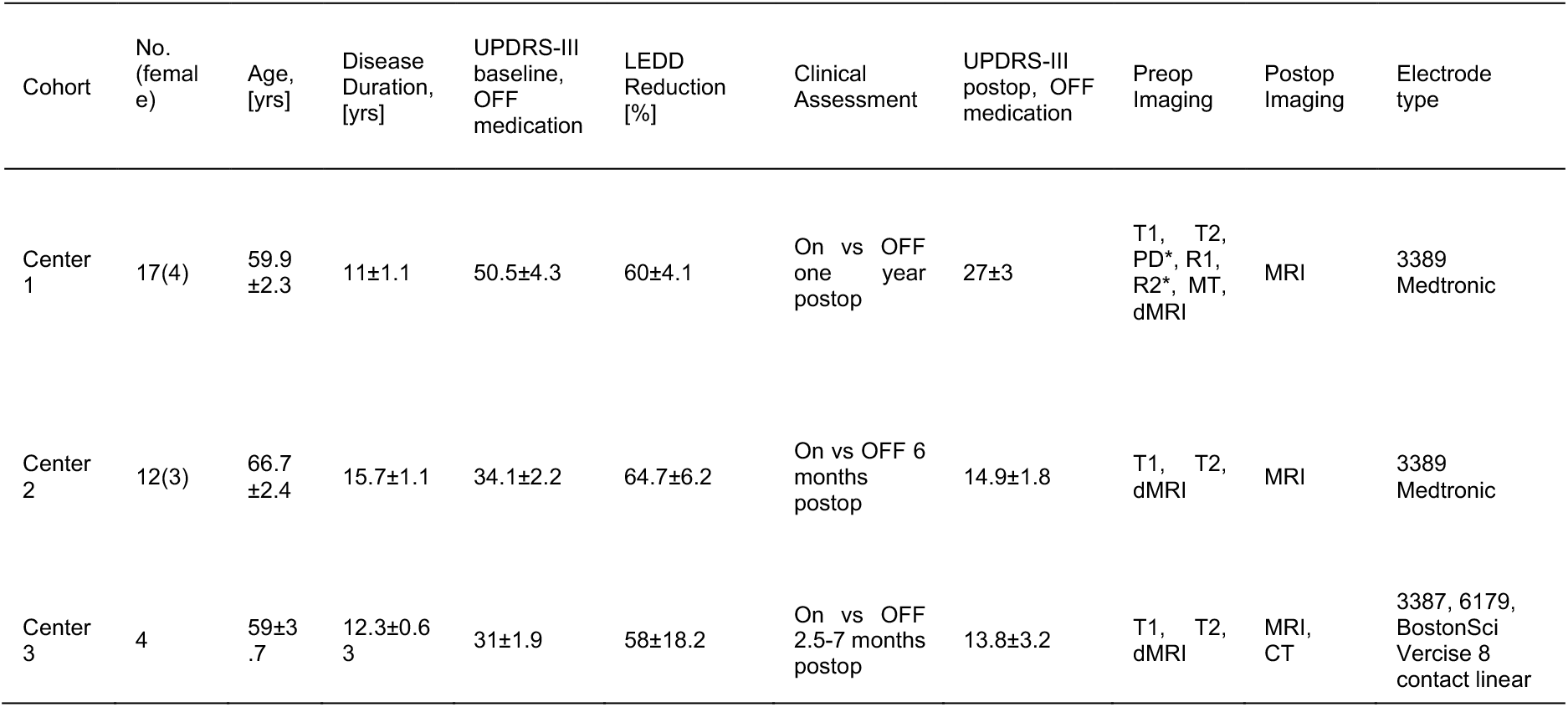
Patient Demographics of All Cohorts Analyzed

All patients underwent stereotactic DBS surgery for treatment of PD and received bilateral DBS electrodes (Table 1). Patients had been enrolled following the standard procedure to screen for eligibility for DBS which excluded structural brain abnormalities or severe psychiatric contraindications. Surgical planning was performed based on MRI imaging and DBS lead localizations were verified by microelectrode recording during surgery and intraoperative macrostimulation for the New York and Mainz centers. Postoperative imaging was carried out to verify accurate electrode placement in all cohorts (see below). Clinical variables including age, sex, disease duration before surgery, L-dopa equivalent dose (LEDD) at baseline were included in analyses. Clinical improvement was measured by comparing Unified Parkinson’s Disease Rating Scale Part III (UPDRS-III) scores OFF medication preoperatively (baseline) and postoperatively ON DBS OFF medication. Improvement was expressed as percentage improvement between the two scores. This study was approved by the local ethics committee of the Charité, University Medicine Berlin (master vote EA2/186/18). The study in London received ethical approval from the West London NHS Research Ethics Committee (10/H0706/68). At Columbia University, all procedures were approved by the Institutional Review Board.

### Preoperative diffusion MRI acquisition

Center 1: For details on the London dataset, please see (Akram et al., 2017). Briefly, imaging data were acquired on a 3 T Siemens Magnetom Trio TIM Syngo MR-B17 using a padded 32-channel receive head coil to reduce discomfort and head motion. Siemens’ 511E-Advanced Echo Planar Imaging Diffusion WIP was used. In-plane acceleration was used (GRAPPA factor of 2) with partial Fourier 6/8. In-plane resolution was 1.5 × 1.5 mm_2_ (Field of view 219 × 219 mm_2_, TR = 12200 ms, TE = 99.6 ms) and 85 slices were acquired with a 1.5 mm thickness. Diffusion-weighting with b = 1500 s/mm_2_ was applied along 128-directions uniformly distributed on the sphere and seven b = 0 s/mm_2_ volumes were also acquired. To correct for distortions all acquisitions were repeated with a reversed phase encoding direction (left to right and right to left phase encode) giving a total of 270 vol acquired ([128 + 7] × 2). The total acquisition time for the DWI sequences was 61 minutes.

Center 2: Diffusion-weighted imaging from Mainz was acquired with 32-directions at b = 1000 s/mm_2_ and one b = 0s volume. DWI of the whole brain at 2 mm isometric voxel resolution covering a field of view of 224×224 mm was obtained. We recorded three acquisitions of DWI sequences encompassing 32 gradient directions and five b0 (no diffusion weighting) images for each acquisition (b value=1000s/mm2, TE=59 ms, TR=11,855 ms, fat saturation “on”, 60 contiguous slices). The total acquisition time for the whole protocol was 35 min which included 24 min (3×8 min) for DWI sequences.

Center 3: The diffusion weighted image sequences from New York cohort were acquired with the following parameters: repetition time: 8500 ms, echo time: 108 ms, slice thickness: 2.50 mm, spacing between slices: 2.50 mm, in-plane resolution 1.88 × 1.88mm_2_, flip angle: 90°, b = 1000 s/mm_2_, 6 b = 0 s/mm_2_ volume, 64 non-collinear gradient directions. To correct for distortions, three acquisitions (2 b = 0 s/mm_2_ and 1 b = 1000 s/mm_2_ volumes) were repeated with a reversed phase encoding direction (left to right and right to left phase encode) giving a total of 73 components ([64 + 6] + 3).

### Diffusion pre-processing and Tractography

For all but the Mainz cohort (where only one b0 volume was acquired), the diffusion (dMRI) data were acquired with reversed phase-encode blips (left-to-right and right-to-left), resulting in pairs of images with distortion going in opposite directions. From these pairs, the susceptibility induced off-resonance field was estimated using a method described by (Andersson et al., 2003) as implemented in FSL (Smith et al., 2004) and the two images were combined into a single corrected one using Topup as implemented in FSL v5.0. The output from Topup was then fed into Eddy (FSL v5.0) for correction of eddy current distortions and subject movement (Andersson and Sotiropoulos, 2016). In the Mainz cohort, only Eddy was applied.

Tractography was done using the generalized Q-sampling imaging method (F. C. Yeh et al., 2010) as implemented in DSI studio (http://dsi-studio.labsolver.org) using the default parameter sets implemented in Lead-Connectome (www.lead-connectome.org; Horn et al., 2014b), which included whole-brain fiber tracking in patient space and transform of the tractogram into ICBM 2009b Nonlinear Asymmetric (‘MNI’) space (Fonov et al., 2011). Whole-brain tractograms were estimated by random-sampling of seedpoints within a white-matter mask that was defined by i) segmenting structural (T1 & T2) imaging data using the New Segment approach as implemented in SPM12 (Ashburner and Friston, 2005) and ii) linearly co-registering the mask to b0-space. In total, 200,000 fiber streamlines were estimated in each patient. Tractograms were then nonlinearly warped into standard space using Advanced Normalization Tools (ANTs; stnava.github.io/ANTs/; Avants et al., 2008) using the “Effective: Low Variance + subcortical refinement” preset implemented in Lead Connectome (Ewert et al., 2019a). Naturally, the same warp was used as in the process of transferring DBS electrodes into standard space (see below).

### Localization of DBS electrodes and VTA Estimation

DBS electrodes were localized using the Lead-DBS toolbox (https://www.lead-dbs.org/; Horn and Kühn, 2015) in updated form (version 2.1.7; Horn et al., 2019) using PaCER (Husch et al., 2018) or the TRAC/CORE approach (Horn and Kühn, 2015) for either postoperative CT or MRI, respectively. Briefly, postoperative CT or MRI were linearly co-registered to preoperative MRI using ANTs. Subcortical refinement was applied as implemented in Lead-DBS to correct for brain shift that may have occurred during surgery. All preoperative volumes were then normalized into MNI space applying the same deformation fields used above.

Based on the long-term DBS settings, volumes of tissue activated (VTA) were estimated using a Finite Element Method (FEM)-based model as implemented in Lead-DBS (Horn et al., 2019). This model estimates the E-field (i.e. the gradient distribution of the electrical charge in space measured in volt per millimeter) on a tetrahedral four-compartment mesh including grey & white matter, electrode contacts, and insulating parts. Grey matter was defined by the DISTAL atlas (Ewert et al., 2018) for relevant structures (STN, internal and external pallidum, red nucleus). The electric field (E-field) distribution was then simulated using an adaptation of the FieldTrip-SimBio pipeline (Vorwerk et al., 2018) that was integrated into Lead-DBS (https://www.mrt.uni-jena.de/simbio/; http://www.fieldtriptoolbox.org/). The E-Field gradient was then thresholded for magnitudes above a commonly used value of 0.2V/mm to define the extent and shape of the VTA (Astrom et al., 2015). Figure 1 provides an overview of the methodology applied.

**Figure 1:**
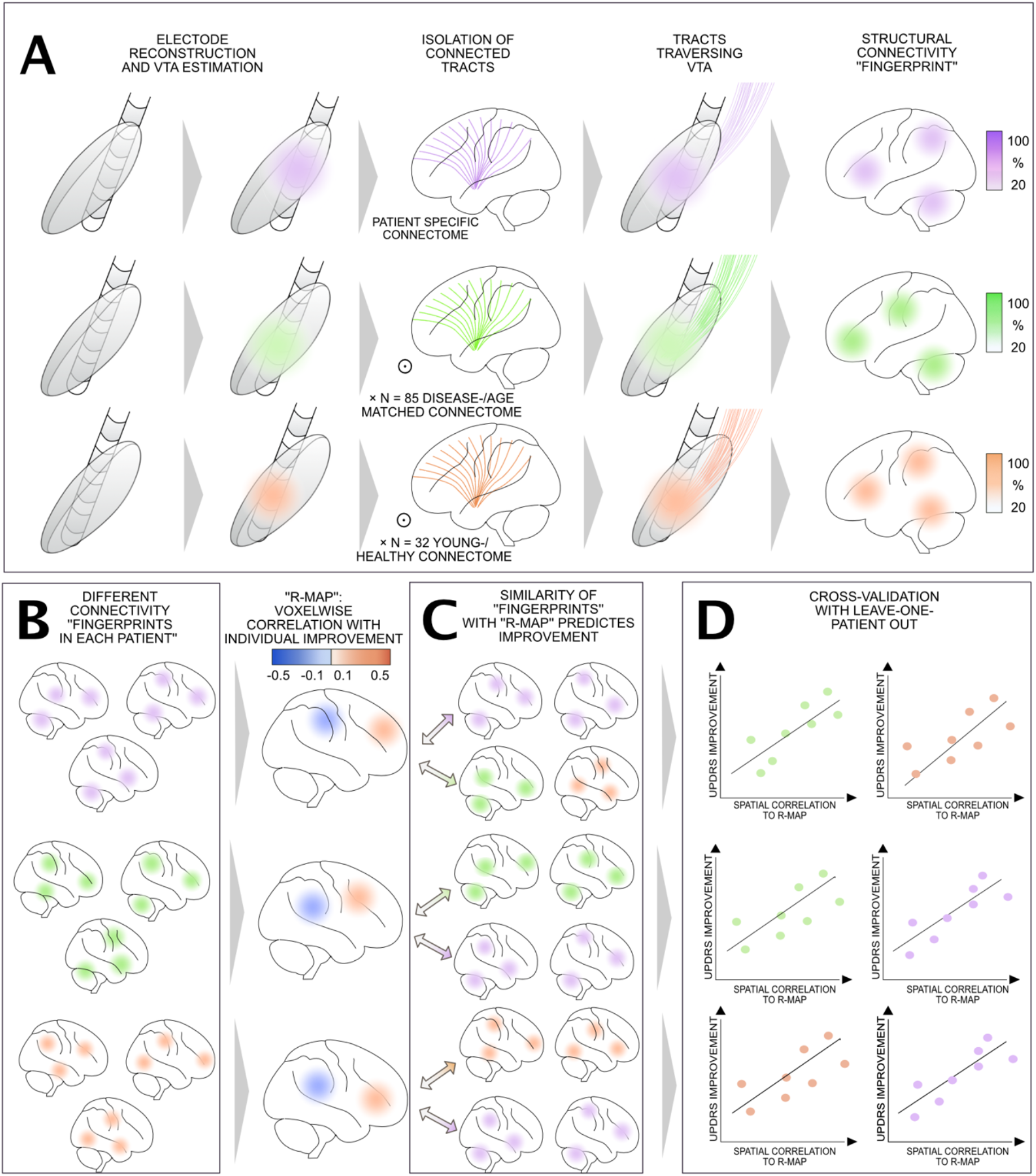
Applied methodological pipeline of data analysis: A) For each patient, DBS leads were localized in MNI space using Lead-DBS software, and volumes of tissue activated (VTA) were estimated based on the actual DBS stimulation parameters. Streamlines representing traversing through each patient’s VTA to the rest of the brain were selected from either patient-specific dMRI data, an age- and disease-matched group connectome or a young-/health group connectome, resulting in DBS stimulation-dependent connectivity “fingerprints”. B) Connectivity “fingerprints” were obtained for each patient using each of the three sources of connectivity data. Across each group of patients, an optimal connectivity profile (R-Map) was generated by correlating connectivity fingerprints with UPDRS-III improvement. C) R-maps represent models for “optimal” connectivity fingerprints. Comparing similarities between each new (out-of-sample) patient’s connectivity fingerprint with these models (by means of spatial correlation), clinical improvements can be estimated (shown in D for a leave-one-out design).

### Structural connectivity estimation

Whole-brain structural connectivity profiles seeding from bilateral VTA for each patient were calculated using three different approaches: First, patient-specific dMRI data were processed for each patient, individually. Second, a disease- and age-matched connectome was estimated based on a cohort of 85 Parkinson’s Disease patients acquired within the Parkinson’s Progression Markers Initiative (PPMI). Third, connectivity profiles were estimated based on a state-of-the-art multi-shell dMRI dataset based on 32 healthy young subjects that were scanned on specialized MRI hardware within the Human Connectome Project (HCP).

### Patient-specific dMRI data

Each patient’s specific connectome was based on dMRI data acquired pre-operatively. Using the generalized q-sampling imaging approach (which was applied in all three modalities) as implemented in DSI Studio (http://dsi-studio.labsolver.org/, F.-C. Yeh et al., 2010), a whole-brain set of 200,000 fiber tracts was estimated using the default processing stream of Lead Connectome (www.lead-connectome.org; Horn and Blankenburg, 2016). This led to one set of (whole-brain) tracts in each patient.

### Age-/ Disease-Matched Connectome

dMRI data from 85 patients were obtained from the Parkinson’s Progression Markers Initiative (PPMI) database (Marek et al., 2011) and processed using DSI Studio / Lead-Connectome in the same fashion as described above. This PPMI connectome of PD patients that is approximately age- and sex-matched to our full cohort was previously computed (Ewert et al., 2018) and has been used in DBS studies, before (Horn et al., 2017b; Irmen et al., 2019; Neumann et al., 2018). Detailed scanning parameters can be found on the project website (www.ppmi-info.org) while processing details can be found in (Ewert et al., 2018).

### Young / Healthy Connectome

dMRI data from 32 healthy young subjects of the Human Connectome Project at Massachusetts General Hospital (https://ida.loni.usc.edu/login.jsp, Setsompop et al., 2013) were obtained and processed using DSI Studio / Lead-Connectome in the same fashion as described above. This ‘MGH Adult Diffusion’ dataset of the HCP was acquired using state-of-the-art scanning sequences on specialized hardware and from a quality perspective may be considered as one of the best openly available, in-vivo dMRI datasets.

Structural connectivity between each VTA and voxels in the rest of the brain was estimated using the above connectome datasets and led to whole-brain fiber-density maps as produced by the Lead Connectome Mapper software (Horn et al., 2019). To do so, fibers traversing through the VTA were selected from the group connectome and projected to volumetric space of the brain in template space of 2 mm isotropic resolution, denoting the number of fibers (connected to the VTA) that traversed through each voxel.

### Estimating a model of optimal connectivity profiles

To estimate a model of optimal connectivity, structural connectivity maps (based on either patient-specific, age- and disease-matched or young/health data), seeding from bilateral VTA were Spearman rank-correlated with %-UPDRS-III change across patients. This led to a map that showed positive or negative associations with UPDRS-III improvements (henceforth referred to as R-maps). Spearman’s correlation was used because structural connectivity results are non-Gaussian distributed (Horn et al., 2014b). These R-maps denote to which areas connectivity is associated with beneficial or detrimental outcomes. In doing so, their spatial distribution estimates the optimal connectivity profile of STN-DBS electrodes for PD that could maximize motor improvements.

### Estimating improvement in out-of-sample patients

To estimate DBS outcome in out-of-sample data, spatial correlations between the optimal structural connectivity model (defined by R-maps or a previously published optimal model from Horn et al. 2017) and the VTA-derived structural connectivity profile in each patient were calculated. For instance, in some analyses, this was done in a leave-one-out fashion (i.e. data from patients #1-32 were used to create an R-map, this map was used to estimate the outcome in patient #33 and so on). This resulting similarity index (again expressed as a Spearman’s rank correlation coefficient) estimates ‘how optimal’ each connectivity profile was and was then used to explore the variability of clinical improvement in a linear regression model.

Throughout the paper, we used randomized permutation tests (5000 permutations) to test for significance (at a 5% significance level) and all analyses were carried out in MATLAB (The Mathworks, Natick, MA).

## Results

Our DBS cohort included 33 patients enrolled at 3 independent centers (7 females, mean age 62.5 ±1.6 years). The average disease duration in the entire sample was 12.9 ±0.8 years. Reduction in LEDD comparing baseline to post-DBS on average was 61.5 ±3.6%. Baseline UPDRS-III score was 42.0 ±2.8, postoperative score 28.1 ±2.1 points (leading to a 51.0 ±3.0% improvement). Total LEDD and UPDRS-III improvements were not significantly different across the three datasets (*p* > 0.05 for all three variables, see table 1 for mean values).

DBS electrode placement was comparable across the three cohorts (Fig. 2) and structural connectivity profiles from two typical patient cases are shown in Figure 3. The visualization of structural connectivity based on the three different connectivity estimates in index patient #1 was almost similar. This was the case for most patients, and fibers predominantly connected to the sensorimotor strip (M1, SMA or pre-SMA). Only in a few patients, patient-individual structural connectivity estimates differed qualitatively, one such example is shown in the bottom row of Figure 3 (index patient #2).

**Figure 2:**
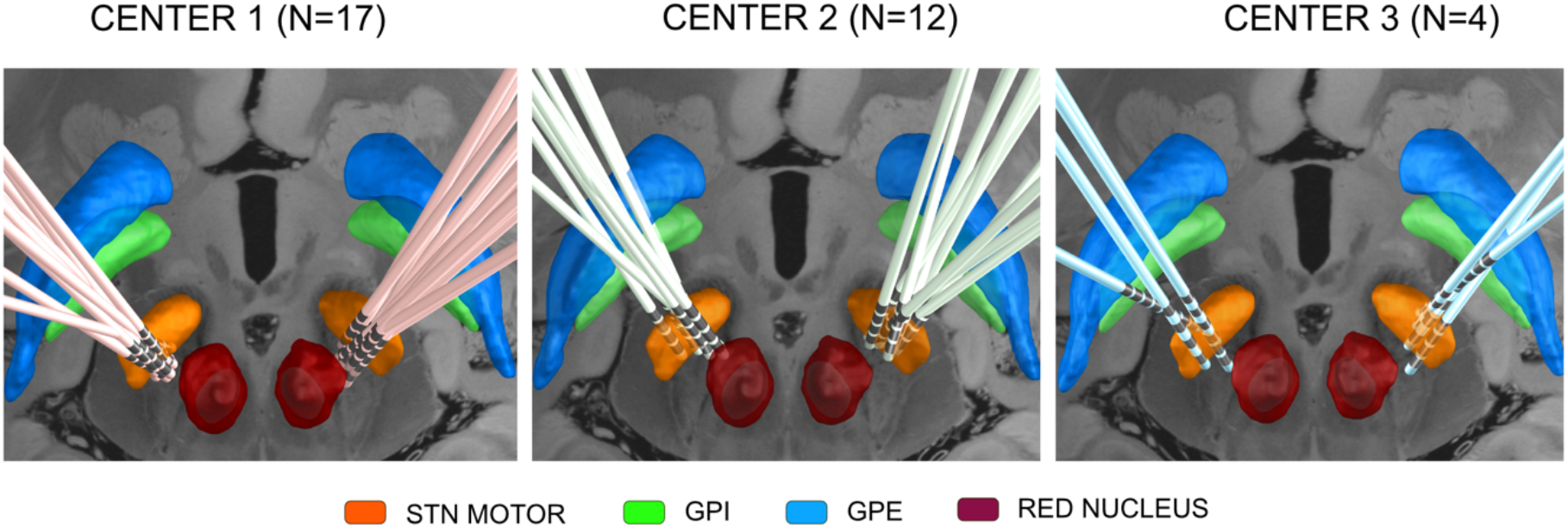
DBS electrode reconstructions in the three cohorts from the three centers. Subcortical structures are defined by the DISTAL Atlas (Ewert et al., 2018), an axial plane of the 7T MRI *ex vivo* human brain template (Edlow et al., 2019) is shown at z = −10 mm.

**Figure 3:**
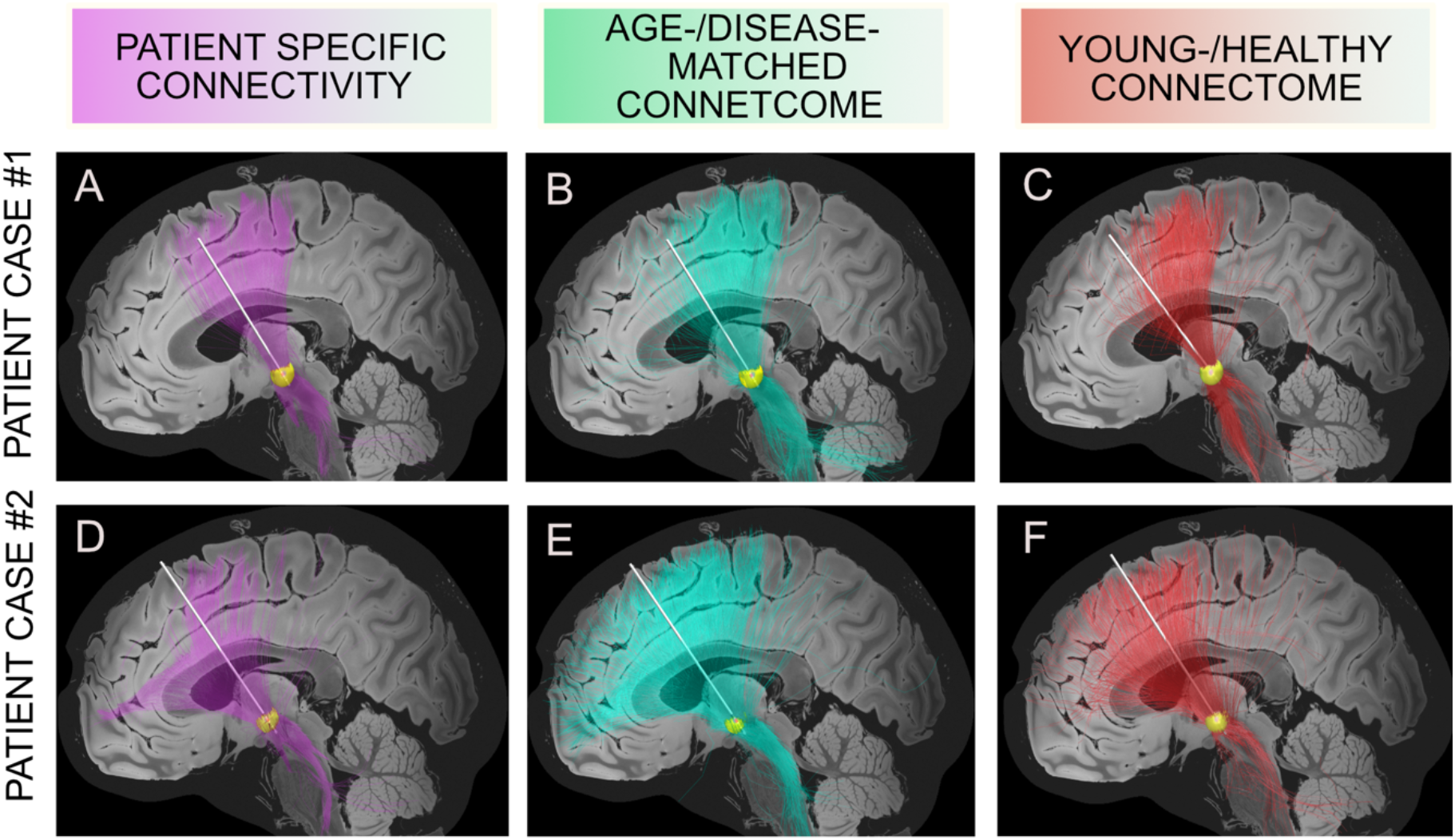
Two representative examples of structural connectivity between individual DBS sites (left hemisphere) and the rest of the brain based on patient-specific connectivity, disease- & age-matched connectome and young-/healthy connectome. A sagittal plane of the 7T MRI *ex vivo* human brain template (Edlow et al., 2019) is shown at × = 3 mm. The yellow sphere represents the VTA.

Correlations between patient-specific and young-/healthy connectivity estimates across the group of patients were high for apical cortical regions but lower for more frontal regions (see Fig. S1). Specifically, connectivity between the electrodes and primary motor cortex was R = 0.57, p = 0.001, supplementary motor area R = 0.40, p= 0.005, pre-supplementary motor area pre-SMA, R = 0.33, p = 0.022 and dorsomedial prefrontal cortex R = 0.21, p = 0.17. Correlations between patient-specific and age-/ disease-matched connectivity, young-/healthy and age-/disease-matched connectivity were shown in Table 2.

**Table 2:**
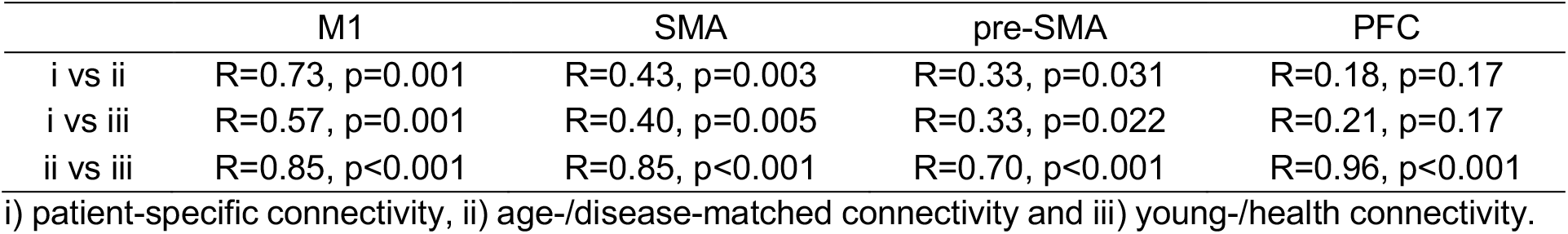
Correlations between connectivity metrics. R-values show agreement between connectivity estimates connecting VTAs to M1, SMA, pre-SMA, and PFC across the group of patients. For instance, the first entry denotes that connectivity strength between VTAs and M1 estimated using patient-specific connectivity (i) and an age-/disease-matched connectome (ii) correlated by a Spearman’s rho of 0.73. Note that similarities between patient-specific and group-level data become lower when advancing in frontal direction while it remains high between normative connectomes.

|  | M1 | SMA | pre-SMA | PFC |
| --- | --- | --- | --- | --- |
| i vs ii | R=0.73, p=0.001 | R=0.43, p=0.003 | R=0.33, p=0.031 | R=0.18, p=0.17 |
| i vs iii | R=0.57, p=0.001 | R=0.40, p=0.005 | R=0.33, p=0.022 | R=0.21, p=0.17 |
| ii vs iii | R=0.85, p<0.001 | R=0.85, p<0.001 | R=0.70, p<0.001 | R=0.96, p<0.001 |

i) patient-specific connectivity, ii) age-/disease-matched connectivity and iii) young-/health connectivity.

In Horn et al. 2017, an R-map that defined optimal connectivity values was estimated based on a two-center cohort of N = 95 patients. In a first step, this R-map was used to account for a certain percentage of outcome in this independent three-center cohort based on individualized diffusion data (Figure 4; R = 0.28, *p* = 0.045). When instead using the age- & disease-matched connectome (R = 0.30, *p* = 0.031) or the young- /healthy connectome (R = 0.33, *p* = 0.021), correlations were also significant. These estimates were not significantly different from each other in head-to-head comparisons based on a Fisher r-to-z transformation (p > 0.8 for all comparisons).

**Figure 4:**
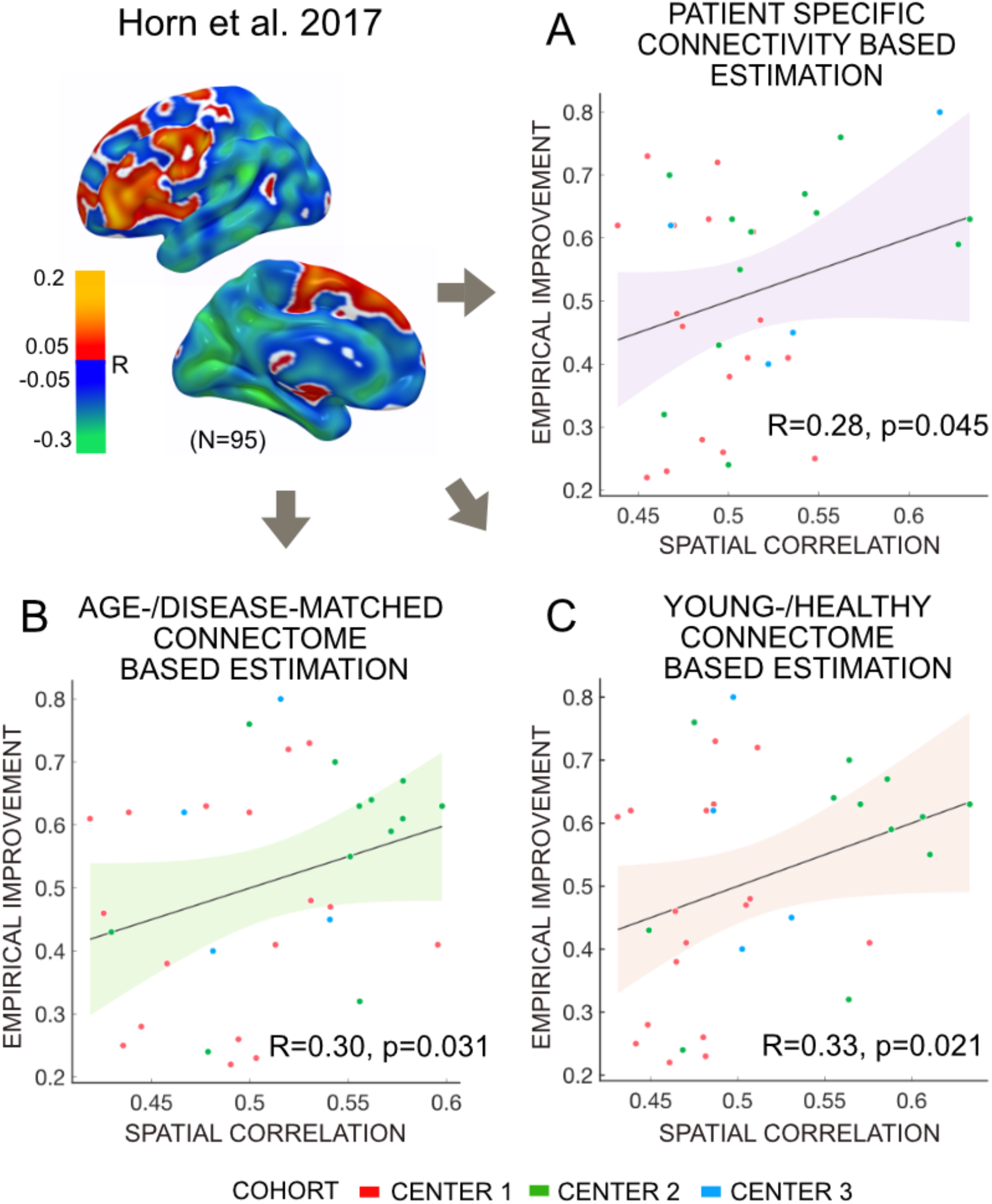
Validation of optimal connectivity profiles estimated in Horn et al. 2017 on the present three-center cohort (N = 33), in which individual dMRI data was available. Clinical improvements could be significantly estimated using patient-specific connectivity (A), a disease-/age-matched connectome (B) and a young-/healthy connectome (C).

In a next step, we calculated data-driven optimal connectivity maps (R-maps) based on the current three-center cohort using patient-specific dMRI, the age-/disease-matched connectome or the young-/healthy connectome, respectively. Using either metric, connectivity to primary motor cortex (M1) and primary somatosensory cortex (S1) was negatively correlated with DBS outcome. In contrast, connectivity to pre-SMA, anterior cingulate and medial frontal cortices was associated with beneficial DBS outcome (Fig. 5).

**Figure 5:**
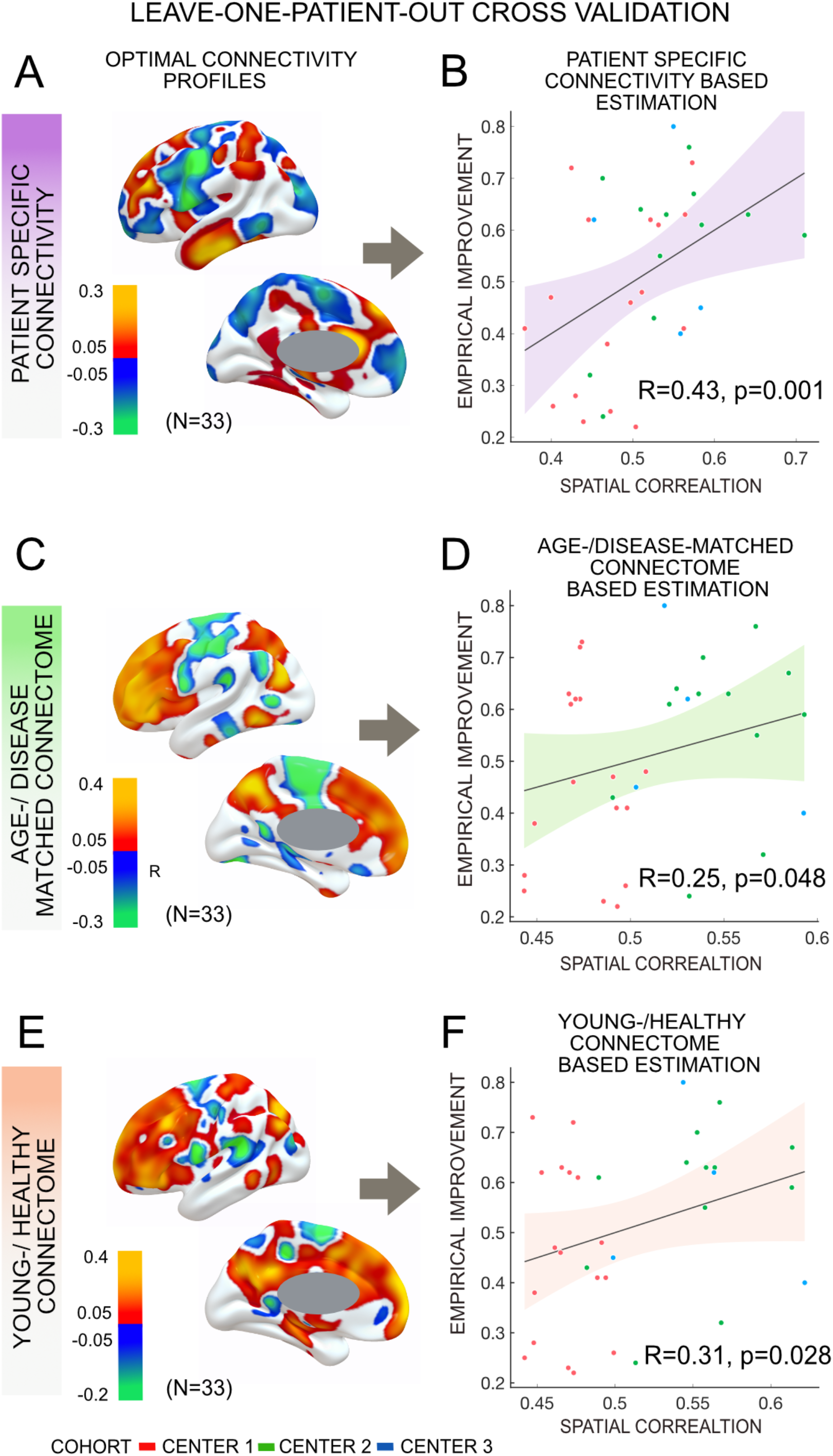
Structural connectivity (patient-specific connectome, age-/disease-matched and healthy-/young) predicted change in UPDRS-III score using a leave-one-patient-out model (N = 33). Optimal structural connectivity model generated with patient-specific connectome (A), age-/disease-matched connectome (C) and young-/healthy connectome (E) effectively estimated patient’s improvement based on respective connectome (B, D, F). Slightly more variance was estimated from the patient-specific connectivity model than the other two metrics.

All three metrics, i.e. patient-specific connectivity (R = 0.43, *p* = 0.001), the age-/disease-matched connectome (R = 0.25, *p* = 0.048) and the healthy-/young connectome (R = 0.31, *p* = 0.028) could significantly account for part of the variance in clinical outcome in a leave-one-out design (Fig. 5). Correlations derived from either metric were not significantly different from each other in head-to-head comparisons based on a Fisher r-to-z transformation (p > 0.4 for all comparisons). Furthermore, cross-estimates between metrics were poorer and not significant, i.e. when the R-map was based on a group connectome, but structural connectivity maps were based on patient-specific structural connectivity or vice versa (Fig. 6). This is not-surprising since normative and patient-specific connectivity estimates are not completely interchangeable. Still, it may underline the importance of consistency in the choice of metric when performing such dMRI based connectivity analyses.

**Figure 6:**
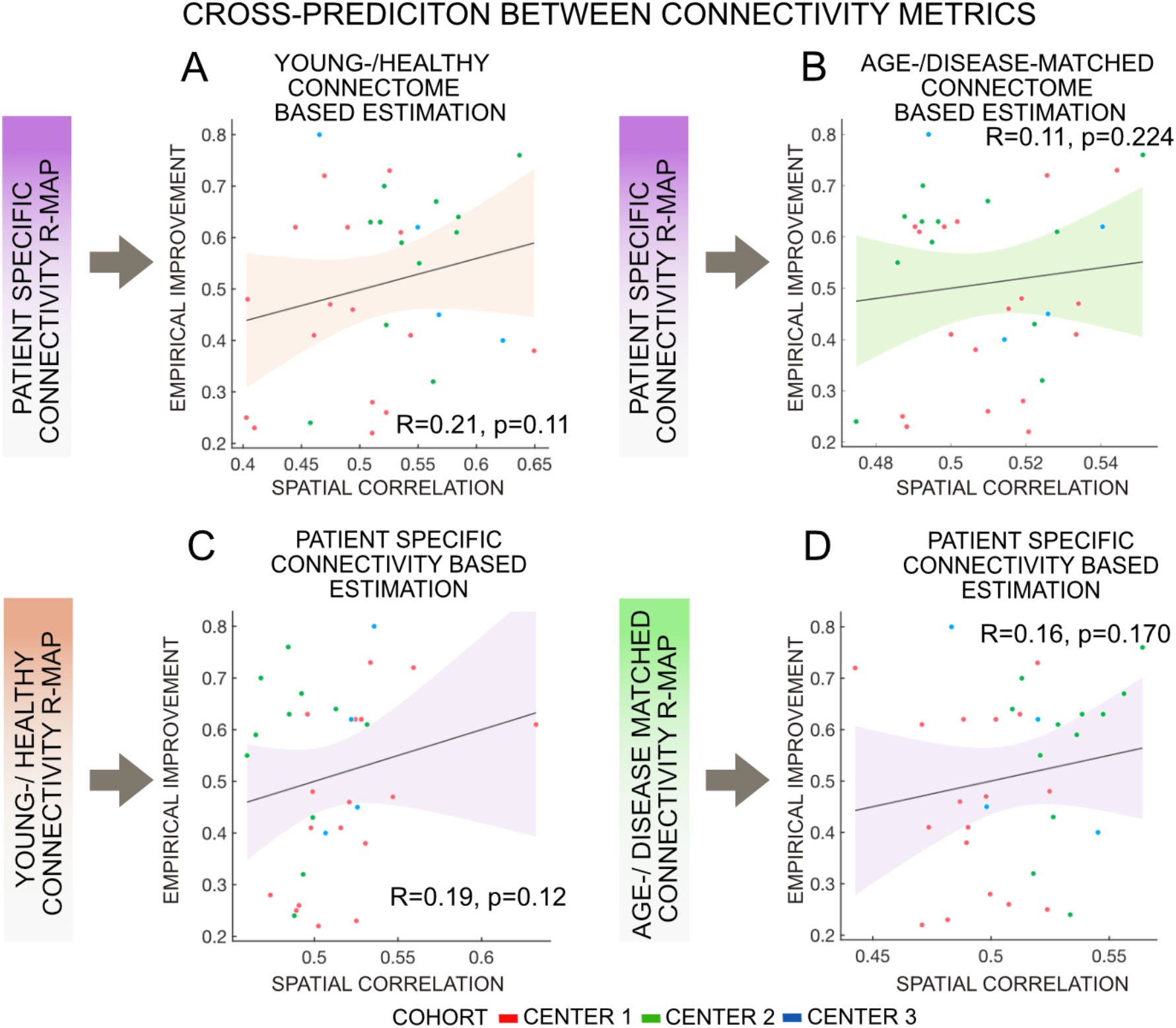
Structural connectivity (patient-specific, age-/disease-matched, healthy-/young connectome) cross-prediction change in UPDRS-III scores under STN-DBS. Patient-specific connectivity couldn’t explain patient’s change following DBS based on healthy-/young connectome (A) and age-/disease-matched connectome (B). Similarly, R-Map generated with young-/healthy connectome (C) and age-/disease-matched connectome (D) couldn’t predict patient’s improvement based on patient-specific connectome.

## Discussion

Three main conclusions may be drawn from the present study. First, the findings support a previously published model of optimal STN-DBS electrode connectivity based on a novel independent sample of patients operated in three different centers (Horn et al., 2017b). Crucially, while the original model had been derived from normative connectivity estimates, patient-specific dMRI data was successfully used to account for DBS outcome in the present cohort. This may further validate the use and concept of applying normative connectome data for the estimation of optimal connectivity models in DBS. Second, we show that in the present cohort, optimal connectivity maps defined using individualized data are highly similar to the ones defined using group connectomes. Specifically, irrespectively of using either patient-specific, age-/disease-matched or healthy-/young connectome data, structural connectivity to pre-SMA, anterior cingulate and medial frontal cortices was associated with beneficial DBS outcome. However, third, variability in clinical improvement made with either method was not completely interchangeable. While none of the metrics resulted in significantly higher predictions than the other two, the use of patient-specific connectome data resulted in the highest R-value between estimated and empirical improvements (R = 0.43 vs. 0.31 or 0.25). This was true although the quality of the patient specific diffusion datasets differed between the three centers resulting in an ‘overall’ poorer data quality than found in normative connectomes.

### Normative group connectomes vs. Patient-specific connectivity

Recently, normative structural connectomes were introduced to account for motor improvement (Horn et al., 2017b) and change of depressive symptoms (Irmen et al., 2019) in Parkinson’s disease patients following STN-DBS. The concept was also applied to DBS in Essential Tremor syndrome (Al-Fatly et al., 2019), Obsessive-Compulsive disorder (J. C. Baldermann et al., 2019; Li et al., 2019) and Epilepsy (Middlebrooks et al., 2018). Furthermore, normative connectomes were used to explain behavioral effects following STN-DBS such as movement speed (Neumann et al., 2018) and motor learning (de Almeida Marcelino et al., 2019). Finally, the concept was applied to investigate side-effects such as DBS induced seizures (Boutet et al., 2019), weight-changes (J. Baldermann et al., 2019) or panic attacks (Elias et al., 2019). While this approach was useful to explore the variability in clinical improvement out-of-sample data (i.e. models were learned on one cohort to account for measures in the other), it was so far not directly compared to the use of patient-specific connectivity.

For instance, in the aforementioned studies, connectivity between the DBS electrodes and the pre-SMA was associated with motor improvement following STN-DBS in PD patients. This was derived by sampling connectivity from the position of DBS electrodes and the average connectivity of that site to the rest of the brain in group connectome datasets. Electrodes situated at a site that was more strongly connected to the SMA yielded significantly better clinical improvement. However, while this may be consistently true when using normative connectomes, it may not hold true when using patient-specific dMRI data. So far, the only study that combined both normative and patient-specific connectivity data in the context of DBS was carried out by Baldermann and colleagues (J. C. Baldermann et al., 2019) in 22 patients suffering from Obsessive Compulsive Disorder. Here, patient-specific data in 10 patients were available but lacked in the remaining 12. When optimal connectivity profiles associated with high clinical improvement were learned based on these 12 patients using a normative connectome, the outcome in the remaining 10 could be predicted by the use of their patient-specific connectivity data (R = 0.7, p = 0.01). The same was true for the opposite case (R = 0.6, p = 0.02). Indirectly, this finding suggested that normative connectomes could be used to define models of optimal connectivity that would remain predictive when applying patient-specific connectivity dataset.

Here, we directly compared patient-specific connectivity estimates to the ones derived from group connectomes (which were either age-/disease-matched or were even acquired in a young-/healthy cohort). We show that optimal connectivity profiles that were associated with good clinical improvement in our sample followed the same overall distribution irrespective of the applied connectivity metric. Namely, functional connectivity with M1 was negatively associated with optimal improvement while more frontal regions (such as SMA, pre-SMA and dorsomedial PFC) were positively associated. Using either method, connectivity profiles were able to account for the variability in clinical improvement in out-of-sample patients (leave-one-out design). Moreover, a previously published optimal connectivity is associated with clinical outcome using either method. This finding is crucial since it shows that optimal profiles could potentially be learned based on large cohorts (and even using normative connectomes) and still applied to patient-specific data. Especially when aggregating large cohorts of DBS patients, it is complicated if not impossible to obtain diffusion-weighted imaging data from them, as well. For instance, large clinical endeavors such as the Early-Stim study cohort (Schuepbach et al., 2013) or the nonmotor study cohort of the International Parkinson and Movement Disorder Society (Dafsari et al., 2018) were acquired without diffusion-weighted imaging data but could still be used to inform optimal connectivity profiles. Similarly, some DBS cohorts are rare to unique world-wide and individualized connectivity data was not acquired for them. Examples include patients suffering from Alzheimer’s Disease stimulated with fornical DBS within the ADvance trials (Laxton et al., 2010), DBS cohorts suffering from rare diseases such as Tourette’s Syndrome (Johnson et al., 2019) or STN-DBS datasets for treatment of cervical dystonia (Ostrem et al., 2011).

### The case for using brain connectivity studies in STN-DBS

Several studies have found significant relationships between electrode placements and clinical outcome, without the need to add connectivity information. Specifically, the same optimal target coordinate within the dorsolateral STN was defined by four independent studies, and three of them showed significant correlations between proximity to this coordinate and resulting clinical improvements (Akram et al., 2017). If such a clear relationship between the local stimulation sites and clinical improvements exist, why should one investigate connectomic mapping at all? The variance explained by such coordinate-based approaches is in the same ballpark of the one explained by brain connectivity. So, what is the added value? We argue that there are several opportunities that motivate the estimation of optimal brain connectivity profiles seeding from DBS electrodes:

Firstly, brain connectivity may bring insights into the mechanism of action of DBS. The concept that strong connectivity to M1 is contra-productive for optimal outcomes but more frontal connections seem favorable qualitatively goes beyond knowledge of an optimal sweet-spot in the STN. From such knowledge, we may derive pathophysiological models and translate findings between systems neuroscience and animal models. Secondly, brain connectivity could at some point be applied to explore the variability of the clinical outcome of novel patients, potentially even before surgery. For instance, work by Muthuraman and colleagues revealed that atrophy in the SMA before surgery was associated with poor clinical outcome following DBS, matching present findings (Muthuraman et al., 2017).

Thirdly, individual patient specific connectivity may differ from the norm and could one day help identify patient-specific DBS targets. In some diseases, where clear associations between clinical outcome and structural tracts have been established, this is already clinical practice (Coenen et al., 2014). Similar work has been theoretically explored using functional MRI (Andersen and Buneo, 2002). So far, to the best of our knowledge, whole-brain connectivity profiles as the ones explored here have not been used in clinical practice, although the general concept has been introduced in 2015 (Fernandes et al., 2015).

Fourthly, networks that lead to side-effects when stimulated could be identified. For instance, Irmen and colleagues recently reported a connectivity profile that was associated with depressive symptoms following STN-DBS in PD (Irmen et al., 2019). Crucially, this network, centered on the left dorsolateral prefrontal cortex, could be reproduced in three international cohorts and was successfully used to cross-predict depressive symptom changes across all cohorts. Such a robust map of a circuit that leads to depressive symptoms could be useful to inform stimulation sites that should be avoided. In Essential Tremor, Al-Fatly and colleagues similarly defined networks that were associated with ataxia and dysarthria (Al-Fatly et al., 2019).

Finally, connectivity profiles could be used to bridge the fields of invasive and noninvasive brain stimulation. In 2014, Fox and colleagues demonstrated that across 14 diseases, the same networks seem to be modulated by both invasive and noninvasive neuromodulation (Fox et al., 2014). In PD, excitatory TMS to M1 and inhibitory TMS to SMA had beneficial effects (the opposite cases did not). If DBS is seen as a functional lesion, this matches optimal connectivity profiles defined here and before.

Thus, it seems sensible to investigate brain connectivity measures in the DBS field for reasons that go beyond finding an optimal target coordinate. Now, the question is which connectivity metric should best be used. We review the advantages of normative vs. patient-specific connectivity data in the following.

### Pros and Cons of normative vs. patient-specific connectivity data

In a number of studies, network targets were identified by using connectivity data that was *not* derived from each individual patient (Akram et al., 2017; Al-Fatly et al., 2019; J. C. Baldermann et al., 2019; Calabrese et al., 2015; Cash et al., 2019; Horn et al., 2017b; Petersen et al., 2019; Weigand et al., 2018). One reason for this is data quality. For instance, Calabrese et al. applied a 200 μm isotropic postmortem scan of the brainstem acquired at 7T to be able to resolve the Wernekinck decussation of the dentatorubrothalamic tract. Weigand et al. applied functional imaging data which was averaged across 1000 subjects, leading to a high signal-to-noise ratio (Thomas Yeo et al., 2011). The structural connectome used in Horn et al. was acquired on a customized Siemens 3T Connectom scanner with multi-shell diffusion-encoding gradients and b-values reaching up to 10,000 s/mm_2_ (Setsompop et al., 2013). Petersen and colleagues abandoned MRI-based connectivity altogether and instead created a realistic tract-atlas based on prior anatomical knowledge because the above acquisition parameters are only obtainable in research settings and it is unfeasible to obtain such data quality during routine preoperative clinical scans. While individualized connectivity data was available in the study by Akram and colleagues, group tractography maps were aggregated across patients.

Furthermore, investigating each patient’s individualized connectivity data is challenging due to poor signal-to-noise and test-retest reliability. This was demonstrated in a study by Petersen and colleagues in which the same subject was scanned ten times. In each, the peak of connectivity to motor-/premotor cortices was identified within the STN (Petersen et al., 2017). Distances across peaks were 0.5 – 1 mm on average. When comparing tensor-based deterministic method vs. an advanced probabilistic method, distances between methods even amounted to an average of 1.4 mm. While this subject was scanned using state-of-the-art methods, test-retest reliability will likely be poorer in clinical datasets acquired in movement disorder patients. A variability of ∼1 mm may seem low at first glance but represents half the distance between two DBS contacts and is in the order of distances across responding DBS patients (Horn et al., 2019). Moreover, the displacement between some runs was found to be of several millimeters, transposing the peak of M1-connectivity from the sensorimotor to the associative functional zone of the STN. If an average displacement of ∼1 mm is introduced by patient-specific connectivity measures, it introduces a significant amount of noise to downstream analyses. This source of variance can be eliminated completely by using group-level or postmortem connectomes, but this comes at the cost of losing individualized connectivity information.

Thus, despite the practical and theoretical advantages of normative connectomes, individualized patient connectivity data is needed to reach a goal of personalized DBS. How could gains of individualized connectivity be combined with the robustness of normative connectomes? One answer would be to scan patients repeatedly and to quantify test-retest reliability. For instance, the midnight scan club endeavor acquired MRI based connectivity data of the same subjects in 12 imaging sessions (Gordon et al., 2017). Doing so in each patient that undergoes DBS surgery is impractical and would be very demanding for patients. However, the dataset was recently used to investigate individualized vs. group-level connectivity-based DBS targets (Greene et al., 2019). Similarly, 45 of the 1200 human connectome project participants were scanned twice to allow for quantification of retest error (Van Essen et al., 2013). Such openly available datasets may be used to investigate the test-retest reliability of individualized patients, while similar data would be needed in patients that actually undergo DBS afterwards.

An additional strategy could be to *integrate* patient-specific and normative connectomes and yield hybrid estimates. Patient-specific connectivity-profiles could be matched to variants that are robustly found within large normative cohorts and thus used to reshape normative connectomes. This concept could be used to reduce the amount of noise in individualized patient acquisitions, but such an approach would require further methodological work and validation studies.

This being said, we should not ignore the fact that all group studies will require co-registrations from the group connectome to the patient or vice-versa. This process will introduce registration bias. New methods for direct DWI registrations are being developed. Meanwhile, image acquisition in single patients and MRI technology are also improving. Thus, while group connectomes may show some advantages currently, but in the future, as the quality and speed of patient specific DWI sequences improve, the indication or need for group average templates may be challenged.

### Limitations

There are several limitations that apply to the current study. First, heterogeneity, such as the differences in the MRI acquisition protocol and assessments of UPDRS-III between the three cohorts should be considered. These differences render the cohort heterogeneous, which should bias results toward non-significance. Also, it may match the heterogeneity of clinical DBS cohorts that are usually aggregated across centers (Schuepbach et al., 2013). Second, inaccuracies in lead localization result from the approach of mapping electrodes into MNI space. To minimize the amount of error introduced by this step, we applied a modern neuroimaging pipeline that was specifically designed for the task at hand. Processing approaches that were designed to reduce error included brain shift correction, multispectral normalization with subcortical refinement steps (Horn et al., 2019) and a phantom-validated electrode localization approach (Husch et al., 2018). The normalization strategy applied here was recently evaluated and led to automatic segmentations of the STN that were nearly as precise as manual expert segmentations (Ewert et al., 2019b). Each step of the pipeline was carefully assessed and corrected if needed. Still, the processing steps include errors that could be further reduced by acquiring data of higher resolution as well as test-retest datasets (see above).

A large limitation that applies to both individualized and normative connectivity mapping can be seen in diffusion MRI in general. Tractography using typical methods on typical diffusion MRI datasets was recently found to include four times the amount of false-positive tracts as true-positive tracts (Maier-Hein et al., 2017). This fundamental problem has led other groups that investigate similar topics to abandon dMRI based tractography altogether and to instead use detailed literature- and expert-based anatomical knowledge (Gunalan et al., 2017; Petersen et al., 2019). Together with poor test-retest reliability outlined above, these issues challenge the overall concept of connectomic DBS. It remains to be seen whether dMRI based tractography may indeed hold up to some of the promises outlined here, in the future (or not).

Further improvements in diffusion imaging, with higher spatial and angular resolution and improved MRI gradients could add to the value of this modality (Jbabdi and Johansen-Berg, 2011; Sotiropoulos et al., 2013). Furthermore, the use of high-resolution postmortem connectome data that is available in submillimeter resolution could be advantageous to employ, as well ((Calabrese et al., 2015) again with the same inherent problem of lacking patient-specificity).

Finally, the optimal connectivity model in the current study was based on full UPDRS-III scores which included multiple symptoms in PD patients, such as tremor, bradykinesia, and rigidity. While the current study was not powered to investigate symptom-specific network fingerprints and addressed a different question, future studies should investigate symptom-specific, mood, cognitive and possibly QoL changes models of optimal connectivity fingerprints.

## Conclusions

Our study analyzed optimal connectivity profiles seeding from STN-DBS electrodes based on patient-specific vs. group-level structural connectivity profiles. We demonstrate that on a group level, results from individualized, age- and disease-matched connectomes and healthy-/young connectomes are comparable but not completely interchangeable. Although differences were not significant, results suggest that individualized structural connectivity has the potential to estimate clinical outcomes following STN-DBS slightly better. Still, the use of normative connectomes seems sensible in cases where individualized connectivity data is lacking.

## Data Availability

Raw data (patient MRI and postoperative CTs) cannot be openly shared because it contains patient information. All code used to analyze data presented in the present manuscript is openly available within the Lead-DBS software suite (https://github.com/netstim/leaddbs; https://www.lead-dbs.org/).

## Acknowledgments

This study was supported by the German Research Foundation (DFG grant SPP2041, “Clinical connectomics: a network approach to deep brain stimulation” to AAK as well as Emmy Noether Grant 410169619 to AH). QW was supported by the China Scholarship Council (CSC).

LZ and HA and supported by a grant from the Brain Research Trust (157806) and the National Institute for Health Research University College London Hospitals Biomedical Research Centre. The Unit of Functional Neurosurgery is supported by the Parkinson’s Appeal and the Sainsbury Monument Trust. The Wellcome Trust Centre for Neuroimaging is supported by core funding from the Wellcome Trust (091593/Z/10/Z).

SG and MM was supported by the German Research Foundation [grant number: CRC-1193-B05, CRC-TR-128-B05, Abbott ISS study-16429 and Boehringer Ingelheim Funds [grant number: BIF-03].

Data collection and sharing for this project were provided by the Human Connectome Project (HCP; principal investigators: Bruce Rosen, MD, PhD, Arthur W. Toga, PhD, Van J. Weeden, MD). HCP funding was provided by the NIH National Institute of Dental and Craniofacial Research, National Institute of Mental Health, and National Institute of Neurological Disorders and Stroke. HCP data are disseminated by the Laboratory of Neuro Imaging at the University of Southern California. HCP is the result of efforts of coinvestigators from the University of Southern California, Martinos Center for Biomedical Imaging at Massachusetts General Hospital, Washington University, and University of Minnesota. Data used in the preparation of this article were obtained from the PPMI database (www.ppmi-info.org/data). For up-to-date information on the study, visit www.ppmi-info.org. PPMI, a public–private partnership, is funded by the Michael J. Fox Foundation for Parkinson’s Research. For funding partners, see www.ppmi-info.org/fundingpartners.

## Supplementary Information

**Figure S1:**
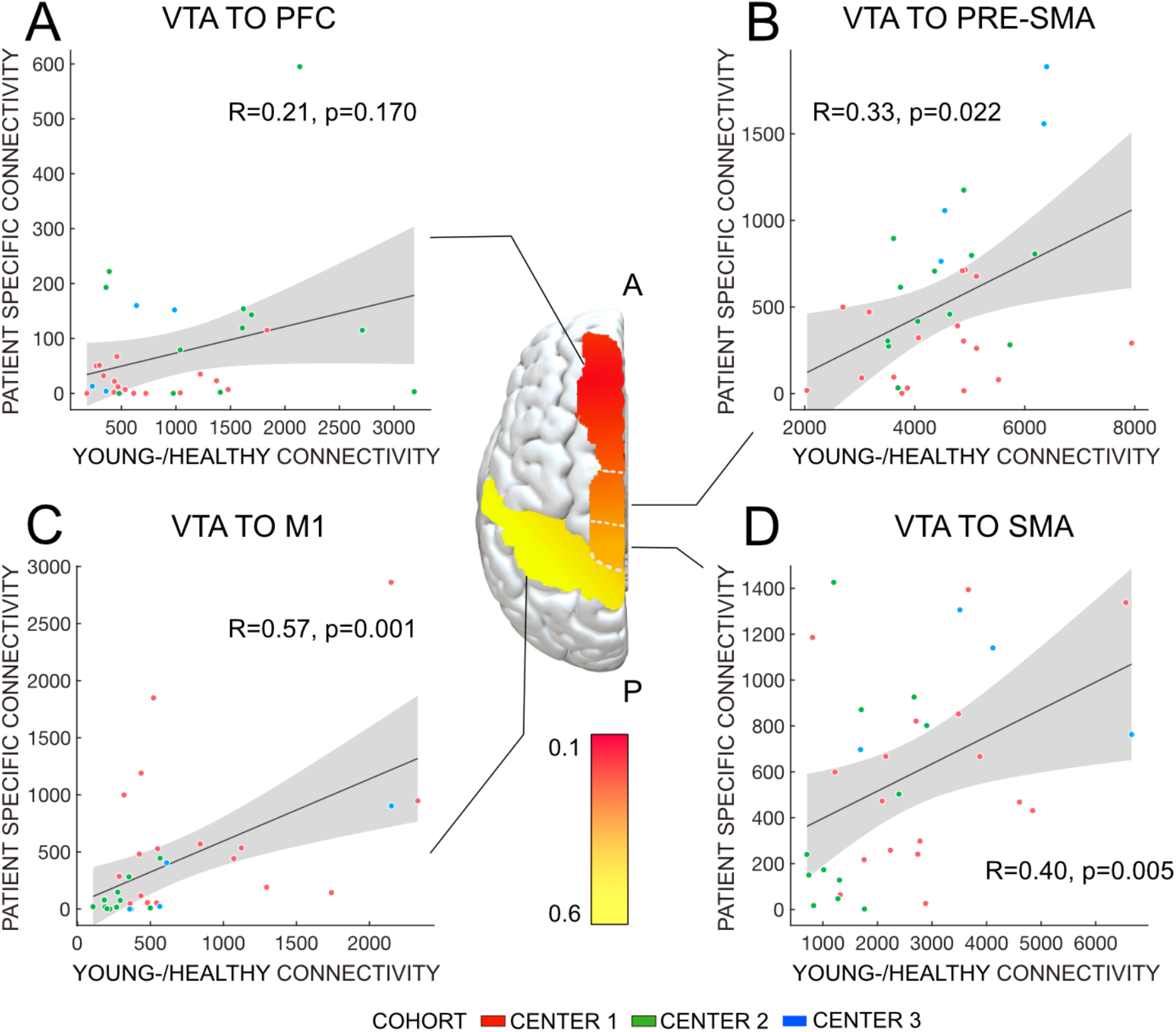
Similarity between patient-specific connectivity and healthy-/young connectivity in M1, SMA, Pre-SMA and dorsomedial PFC in the three cohorts. The similarity degree in M1>SMA>Pre-SMA>dorsomedial PFC.

**Figure S2:**
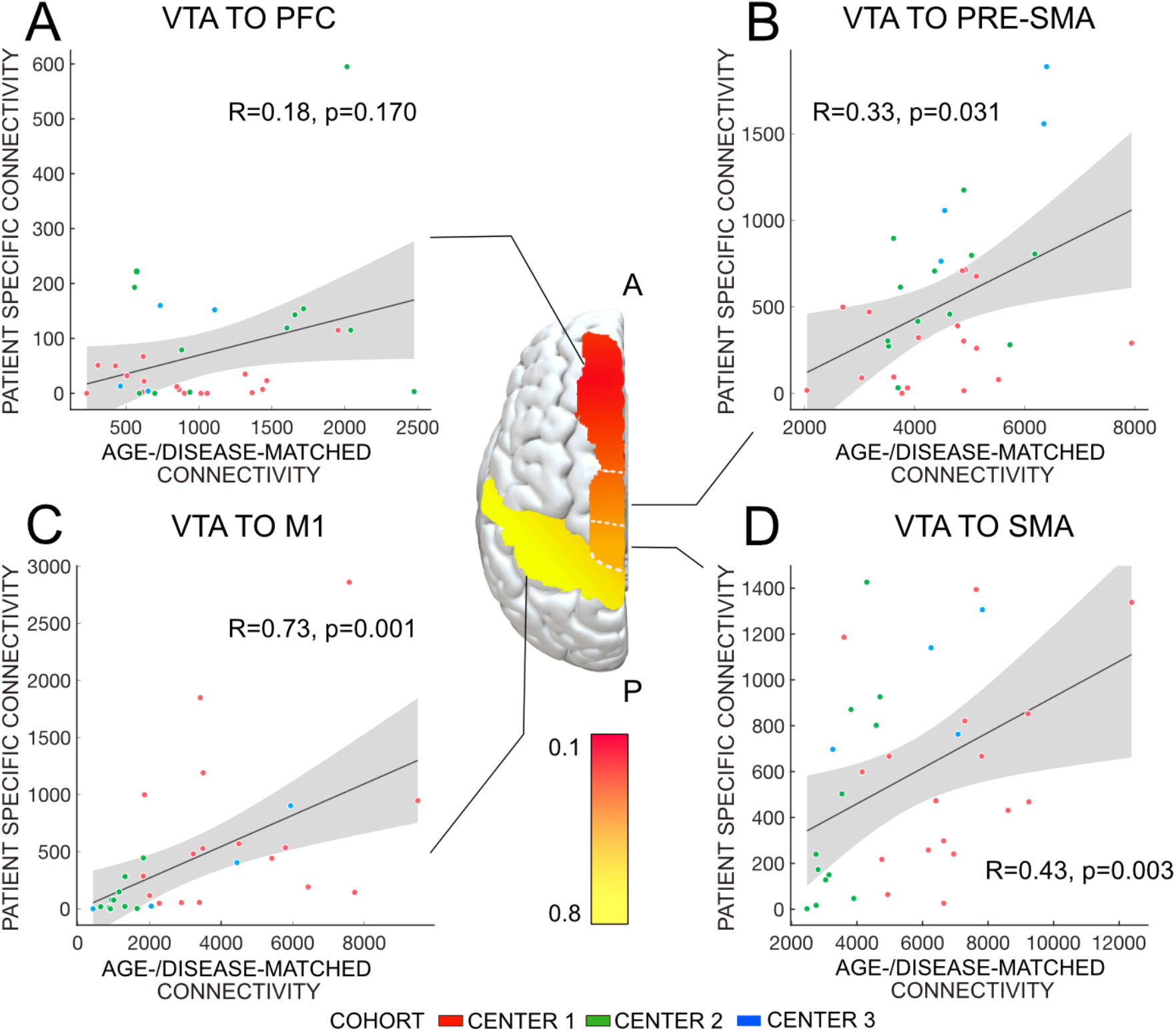
Similarity between patient-specific connectivity and age-/disease-matched connectivity in M1, SMA, Pre-SMA and dorsomedial PFC in the three cohorts. The similarity degree in M1>SMA>Pre-SMA>dorsomedial PFC.

